# Estimation of the severeness rate, death rate, household attack rate and the total number of COVID-19 cases based on 16 115 Polish surveillance records

**DOI:** 10.1101/2020.10.29.20222513

**Authors:** The MOCOS International Research Group, Barbara Adamik, Marek Bawiec, Viktor Bezborodov, Przemyslaw Biecek, Wolfgang Bock, Marcin Bodych, Jan Pablo Burgard, Tyll Krueger, Agata Migalska, MOCOS, Tomasz Ożański, Barbara Pabjan, Magdalena Rosińska, Malgorzata Sadkowska-Todys, Piotr Sobczyk, Ewa Szczurek

## Abstract

**Background:** Estimating the actual number of COVID-19 infections is crucial for steering through the COVID-19 pandemic crisis. It is, however, notoriously difficult, as many cases have no or only mild symptoms. Surveillance data for in-household secondary infections offers unbiased samples for COVID-19 prevalence estimation.

**Methods:** We analyse 16 115 Polish surveillance records to obtain key figures of the COVID-19 pandemic. We propose conservative upper and lower bound estimators for the number of SARS-CoV-2 infections. Further, we estimate age-dependent bounds on the severe case rate, death rate, and the in-household attack rate.

**Results:** By maximum likelihood estimates, the total number of COVID-19 cases in Poland as of July 22nd, 2020, is at most around 13 times larger and at least 1.6 times larger than the recorded number. The lower bound on the severeness rate ranges between 0.2% for the 0–39 year-old to 5.7% for older than 80, while the upper bound is between 2.6% and 34.1%. The lower bound on the death rate is between 0.04% for the age group 40–59 to 1.34% for the oldest. Overall, the severeness and death rates grow exponentially with age. The in-household attack ratio is 8.18% for the youngest group and 16.88% for the oldest.

**Conclusions:** The proposed approach derives highly relevant figures on the COVID-19 pandemic from routine surveillance data, under assumption that household members of detected infected are tested and all severe cases are diagnosed.

**MOCOS:** The MOCOS (**MO**delling **CO**rona **S**pread) international research group is an interdisciplinary scientific consortium. The following authors are MOCOS members: Barbara Adamik, Marek Bawiec, Viktor Bezborodov, Przemyslaw Biecek, Wolfgang Bock, Marcin Bodych, Jan Pablo Burgard, Tyll Krueger, Agata Migalska, Tomasz Ożański, Barbara Pabjan, Magdalena Rosińska, Piotr Sobczyk and Ewa Szczurek

## 1 Introduction

The COVID-19 pandemic has led to dramatic changes in the everyday life worldwide. The most appropriate countermeasures are sought against a further spread. To this aim, key disease characteristics need to be described, such as the severeness and attack rates. Mounting evidence indicates that many infected with SARS-CoV-2 show no or only mild symptoms and remain undetected.^1,2,3^ The lack of accurate knowledge of the actual number of infected highly complicates epidemic control.^4^

Computational approaches to estimating both severity of COVID-19 disease and the unknown total number of infected range from statistical and machine learning to model-based.^6,6,7,8,9,10,11^ With more detailed data, these approaches gain more statistical power and need to make less educated guesses. The report by Bi et al. proved contact tracing and surveillance data to be useful in characterizing epidemiology and transmission of SARS-CoV-2 in China.^5^ To our knowledge, such detailed case data has not been reported in Europe, and has not been used for the estimation of the total number of cases.

Here, we rely on detailed COVID-19 surveillance data in Poland, recording the source of transmission, household contacts and hospitalization status for 16 115 cases. These data allow to differentiate from the cases that were first detected in each household from the secondary cases that were infected in the household. While the former cases are often diagnosed based on their symptoms and are biased towards worse disease progression, the latter can be considered unbiased samples of the susceptible population. Using this unbiased sample, we derive upper and lower bounds on the age dependent rates of severe progression, death rates and in-household attack rates. We show that the severeness rates grow exponentially with age. Unlike easily obtainable case fatality rates, our bounds on death rates give insight into the actual lethality of COVID-19. Based on these estimates, we further provide bounds on the total number of SARS-CoV-2 cases. Our results pave the way for utilizing surveillance data in the COVID-19 spread for unbiased estimation of its key characteristics and the unknown total number of infections.

## 2 Methods

In the following, we analyze COVID-19 surveillance data, with each case assigned to a household. For each household, the first detected case in this household is referred to as an *index case*. The index cases may be subject to some bias, since they may be diagnosed due to the severity of their disease, different kinds of ex-ante social activity or other exposure factors. All remaining household members (all apart from the index cases) are referred to as the *susceptibles*. The number of susceptibles is equivalent to the sum of the household sizes reduced by the number of the index cases, and is denoted *N*^*^. In contrast to the index cases, it is plausible that the infections among the susceptibles are unbiased with regard to clinical progression, severeness rate and attack rate. Those remaining infections in the household are called *secondary household infections*. Let *I*^*^ denote the number of the known (recorded in the data) secondary household infections. Denote by *T*^*^ the true total prevalence of secondary household infections (including undetected cases). Let *I*^*,sev^ be the number of severe cases among these infections, and assume that all severe cases among the susceptibles are observed.

There is a close relation between the rate of severe disease progression, household attack rate and the undiagnosed fraction of COVID-19 infections, as they can all be estimated using the above-mentioned quantities. Below, we first derive estimators for the bounds on COVID-19 severeness and attack rates, as well as the total number of infected, without taking account for such factors as age or sex. Next, we extend the derivation to consider these factors. We considered two alternative definitions of severe cases. First, we assumed severe cases are those hospitalized for 10 days and more or dead, and second we assumed they were hospitalized for 14 days and more or dead. Finally, we describe how the susceptible population size can be estimated from external census data in the absence of records of household sizes for the index cases.

### 2.1 Estimating the severeness rate and the death rate

Given the true total number of secondary household infections *T*^*^ and the number of severe secondary infections *I*^*,sev^, the severeness rate is easily estimated as

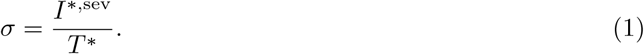

*I*^*,sev^ can easily be obtained from the existing data records, since severe cases are likely found in a functioning healthcare system. Contrarily, e.g. due to asymptomatic cases, *T*^*^ may be unknown. We hence derive upper and lower bounds on the severeness rate, which are derivable from the observed quantities. First, denote the observed severe case rate *α* by

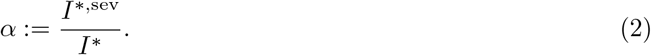

If all infected persons were diagnosed, *I*^*^ = *T*^*^ and *α* would be the true severe case rate among the infected. In common case, when only some of the infected are diagnosed, *I*^*^ is the minimum of secondary infected in the observed households. Hence, *α* defines an upper bound on the severe case rate.

This rate is lower bounded by

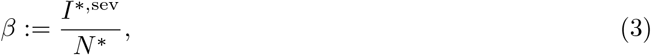

as *N*^*^ is the maximum number of possible infected in the observed households.

In Section 2.3 we compute different bounds on severeness rates, separately for different values of other factors, such as age. Finally, smooth estimates on the functional dependence of the severeness bounds on the age are obtained using restricted cubic splines. Further details are depicted in Section A of the supplementary online material (SOM).

The exact same equations as for the bounds on the severeness rates can be used to estimate the death rates, but inserting for *I*^sev^ the number of deaths instead of the number of severe cases.

### 2.2 Estimating the total cumulative number of infections

Let *T* ^sev^ be the total number of severe cases in the entire population and *T* be the unknown true total number of SARS-CoV-2 cases. Assuming that the severeness rate among secondary infected individuals, *σ*, is the same as it is in the entire infected population, we have

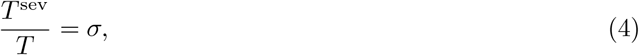

from which we obtain the estimate of the total prevalence *T*.

Since *σ* is often not directly obtainable from the data, we can use it’s lower bound *β* and upper bound *α* to derive the conservative maximum likelihood upper and lower bounds on *T*, respectively as

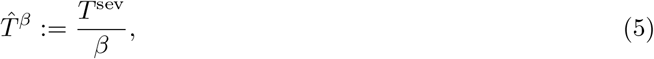

and

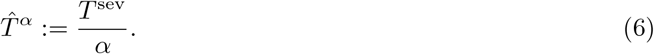

If the secondary infections in the households are diagnosed precisely, this lower bound (6) will be near the expected number of infected. A gap between officially recorded infections and the estimated lower bound could stem from a poor testing of secondary infections. If this can be ruled out, the gap indicates the expected minimum of additional undiagnosed infections.

Conservative one-sided 1% lower confidence bound on *β* (denoted *<u>β_q_</u>*), and 99% upper confidence bound on *α* (denoted 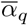), are derived using and extension of the Clopper-Pearson interval method^12^ (SOM B). Based on this, the one-sided 99% upper confidence interval bound for the upper bound of infected is given by

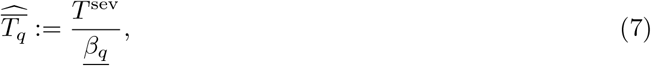

while he one-sided 1% lower confidence interval bound for the lower bound estimator of infected is given by

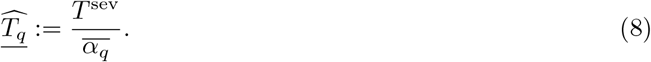

For a rapidly growing number of infections, which was not the case of the Polish epidemic as of July 22nd, 2020, one would have to adjust for a time delay from infection to the recording of severe cases. In SOM C we describe how to correct for this time lag.

### 2.3 Accounting for population strata to estimate of the total number of infections

Above we assumed that the rate of severe disease progression among infected individuals is the same. It is known, however, that this rate depends on factors like age, sex, and the comorbidity status.^9,13,14,15^ When stratifying the population according to these factors, the between-class variance is removed from the total variance. Hence, the estimate becomes more efficient. We thus adapt the approximation to account for these strata. Again, we make use of the fact that the secondary infected in the households constitute a severeness-rate-unbiased sample. Instead of calculating the rates *α* and *β* over all units in the sample, they are calculated in the classes known to affect the severity of infections.

Let us number the classes of all combination of age, sex, comorbidity values consecutively with *l* = 1, …, *L*. Note that the resulting classes, as in ANOVAs, have to be big enough such that some severe cases 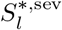 in the *l*-th class are observed. The severe case rates in class *l* yield

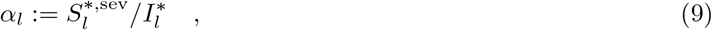

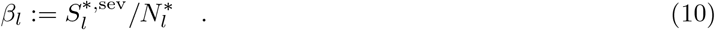

Therefore, for obtaining an upper bound estimator of the upper bound of infected we sum up this figure over all classes.

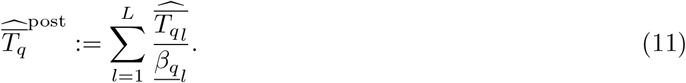

Accordingly, the estimators 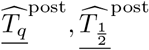, and 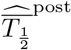 can be obtained.

### 2.4 Estimating the unknown number of susceptibles

In Poland and other countries, neither the negative tests nor the household sizes are recorded. In that case it is necessary and possible to estimate *N*^*^ based on external data. Here, we obtain the household size distribution conditioned on demographic characteristics of the index cases from census data. Via this information a distribution of *N*^*^ is derived by bootstrapping, as described in SOM D. The bootstrap draw of a household size was conditioned on the age of an index case, minimal household size information and residing voyvodship. Adjustment for the estimation of the confidence bound for the lower bound *β* on the severeness rate is described in SOM B.

### 2.5 Estimating the household attack rate

There are two natural ways to define the household attack rate. The first is a probability Λ^*^ of an infected household member to infect a non-infected household member. We refer to this quantity as the *attack probability*. Λ^*^ is estimated based on the secondary case data, taking into account the fact that the infections proceed consecutively within a household (SOM E). The second is the ratio of the expected total number of infections *T*^*^ to the expected number of susceptibles *N*^*^, given Λ^*^ and the distribution of the household size (SOM E). We refer to this quantity as the *attack ratio* and denote it *G*(Λ^*^). Arguably, in contrast to the attack probability, the attack ratio is not a medically-relevant intrinsic characteristic of the disease. It is, in particular, dependent on the household size distribution. Thus, for example, for the same attack probability, the attack ratio would be different for countries with different typical household size. An estimator of the attack ratio is then given by:

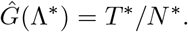

Since the true number of secondary infections *T*^*^ is unknown, as we instead compute the lower bound on the attack ratio as

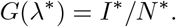

*G*(*λ*^*^) depends on the probability *λ*^*^, which in turn constitutes a lower bound on Λ^*^.

Since the upper and lower bounds on the severeness rates depend directly on the *I*^*^ and *N*^*^, respectively, one can associate attack probability *λ*^*^ directly with the upper bound *α* on the severeness rate, and an attack probability 1 with the lower bound *β*. Furthermore the relation *α ·G* (*λ*\*) = *β* between attack rate, rate of severe progression and the *G* - function holds.

Analog computations can be performed for age-dependent attack probability and ratio (SOM E). In these computations, we assume that the probability and ratio depend on the age of the susceptible (the person acquiring the disease).

### 2.6 Collection of surveillance data

The analyzed data was collected as part of routine COVID-19 surveillance in Poland, which was implemented based on a data collection system functioning for other notifiable infections. The mandatory reporting was ordered both for clinical diagnoses of COVID-19 and positive laboratory tests of SARS-CoV-2. The notifications were sent to the local public health departments, which were responsible for conducting epidemiological investigation, contact tracing and, if necessary, - ordering quarantine.

According to the protocol, all quarantined cases were tested in case of symptoms. Testing of all individuals in the quarantine was optionally applied. The results of the epidemiological investigations were documented in the Epidemiological Reports Registration System (SRWE). The data was to be updated once the case outcome was known. However, given the strain on the public health system, this information could be missing or delayed. The SRWE database includes basic demographic and clinical information, exposure category, hospitalization history and use of mechanical ventilation and moreover detailed information on established links between cases.

### 2.7 Data pre-processing and estimating crucial quantities from surveillance data

The full dataset of 17 359 surveillance cases was pre-processed to leave only records with clear epidemiological links registered (SOM F). The most important predictor for the progression of COVID-19 is the age, so we focus on this variable for creating the classes according to Equation 11.

The total number *T* ^sev^ of the severe cases the Polish population was computed as the number of all such defined severe cases in the analyzed households (including the index cases), multiplied by a factor *d/c*, where *d* was defined as the total number of officially diagnosed cases in Poland (as of July 22nd, 2020), and *c* the total number of all diagnosed in the analyzed database 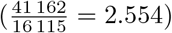. The factor is intended to scale up the database level estimates to a national level of the entire Polish population.

## 3 Results

### 3.1 Surveillance data characteristics

We characterized a total of 16 123 COVID-19 surveillance records (Table 1), out of which 11 895 (73.8 %) were the index cases and 4228 (26.2 %) were the secondary cases. The patients were divided into four age groups, including a group of 0–39 years old (39.8 % of all records). This wide age group was formed to reliably estimate per-group severe case rate, as there were no or only a few severe cases among children. The proportion of females was slightly larger (51.6%) than of males, and similar in both index cases (51.1%) and secondary cases (52.9%).

**Table 1:** Demographic and clinical characteristics of analyzed COVID-19 surveillance dataset including all cases, index cases for household transmission and secondary cases. The percentages in the brackets are of the total number of 16 115 analyzed cases.

|  | All cases<br>no. (%) | Index cases<br>no. (%) | Secondary cases<br>no. (%) |
| --- | --- | --- | --- |
| Total | 16 115 (100.0 %) | 11 888 (73.8 %) | 4227 (26.2 %) |
| Within age group |  |  |  |
| Age 0 - 39 | 6400 (39.7 %) | 4232 (35.6 %) | 2168 (51.3 %) |
| Age 40 - 59 | 6084 (37.8 %) | 4918 (41.4 %) | 1166 (27.6 %) |
| Age 60 - 79 | 2900 (18.0 %) | 2206 (18.5 %) | 694 (16.4 %) |
| Age 80+ | 708 (4.4 %) | 532 (4.5 %) | 176 (4.2 %) |
| Unknown age | 23 (0.1 %) | 0 (0.0 %) | 23 (0.5 %) |
| Sex |  |  |  |
| Female | 8322 (51.6 %) | 6084 (51.2 %) | 2238 (52.9 %) |
| Male | 7793 (48.4 %) | 5804 (48.8 %) | 1989 (47.1 %) |
| Hospitalization |  |  |  |
| Hospitalized | 4373 (27.1 %) | 3602 (30.3 %) | 771 (18.2 %) |
| Hospitalized $\geq 10$ days | 2399 (14.9 %) | 1978 (16.6 %) | 421 (10.0 %) |
| Hospitalized $\geq 14$ days | 1811 (11.2 %) | 1505 (12.7 %) | 306 (7.2 %) |
| Final outcome |  |  |  |
| Deceased | 488 (3.0 %) | 455 (3.8 %) | 33 (0.8 %) |
| Recovered | 5872 (36.4 %) | 4246 (35.7 %) | 1626 (38.5 %) |

The index cases can be regarded as detected based on their symptoms and the secondary cases as an severeness-rate-unbiased sample of the population. The index cases were more often hospitalized (with hospitalization rate 30.3 %) than the secondary cases (18.2 %). In addition a larger fraction of hospitalization for longer than 14 days is observed (12.7 % for index cases vs 7.2 % for secondary cases). On 22/07/2020, the final outcomes were known for 6 360 out of 16 123 cases, with 488 deceased and 5872 recovered. Again, for the index cases, the death fraction was larger than for the secondary cases.

### 3.2 Estimation of the bounds on severe progression and death rates

To estimate upper and lower bounds for the COVID-19 severeness rate in Poland, we focused on the secondary case data (Table 2). The expected number of susceptibles *N*^*^, i.e., the estimate for the number of all inhabitants of the analyzed households, except for the index cases, was computed using the Polish census data (SOM D) as 39 102. First, we obtained the bounds on the severeness rate for the different age groups using the maximum likelihood estimators, in Equation 3 for the lower bound, and in Equation 2 for the upper bound, respectively. For the definition of severity, we used two alternative thresholds of hospitalization, 10 and 14 days respectively. For the 14 days threshold, the young population (0-39 year old) had the smallest lower bound on severe case rate (0.2%). The bound increases for older age groups, reaching 5.7% for the group older than 80. Across the age groups, the upper bound on the severeness rate *α* is roughly an order of magnitude larger than the lower bound *β* and also increases for older age groups.

**Table 2:**
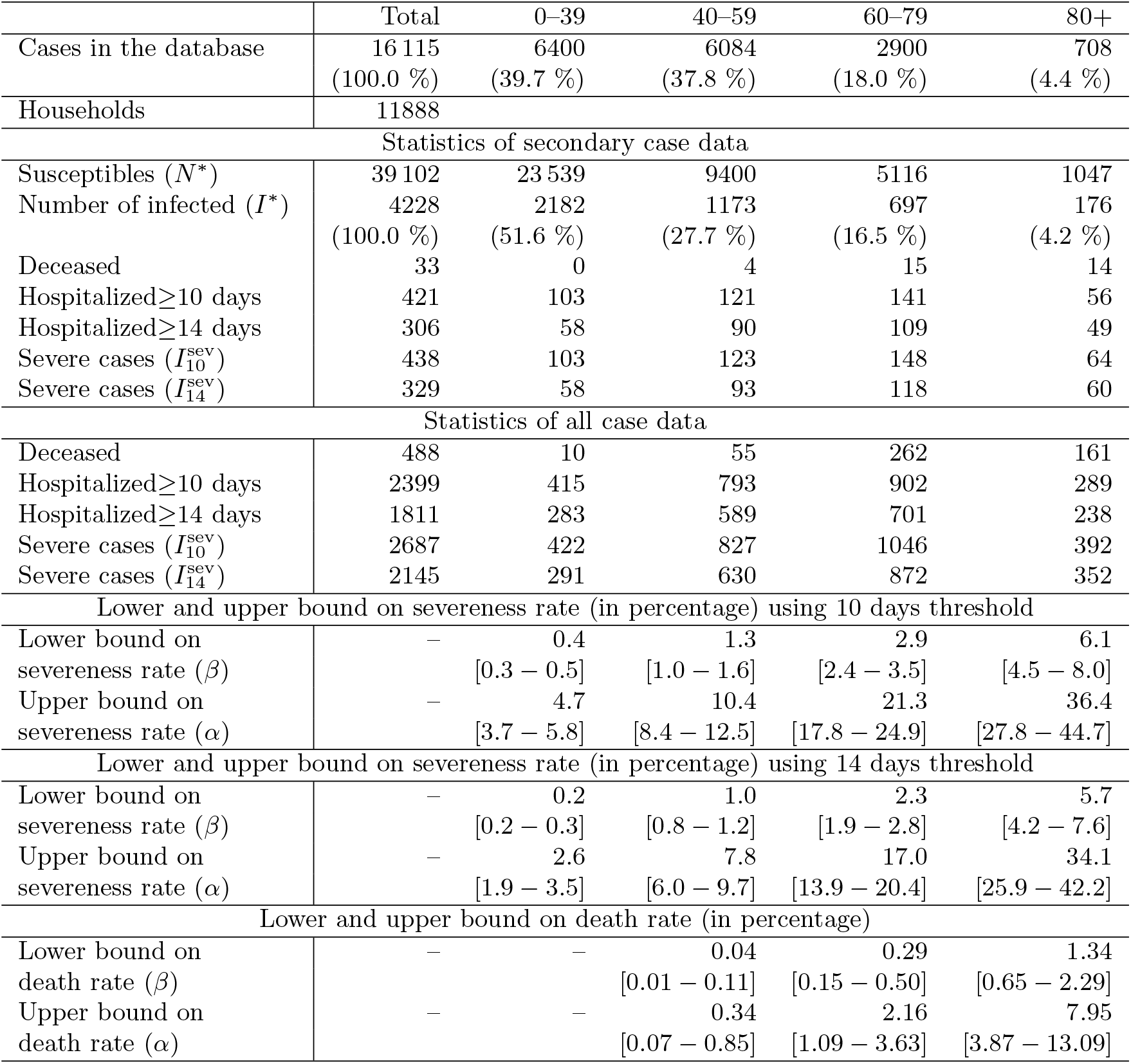
Observed and estimated figures for the COVID-19 pandemic in Poland based on the available database (as of 22/07/2020)

Next, we obtained smooth estimates of the severity rates and death rates among secondary cases as a function of the age together with the 98% bootstrap intervals (Figure 1). The death rate is presented only for people over 60 years, because number of deceased among secondary cases among younger people was too small to get credible intervals. To get estimates at this resolution we estimated mortality with logistic regression model with age transformed with tail linear restricted cubic splines (SOM A). Overall, severity and mortality depend exponentially on the age (note Figure 1 is a semilog-plot). This finding is in agreement with the fact that severe cases are more likely among the elderly patients.^16^ The estimate for *β* is roughly 10x higher than the estimate for *α*.

**Figure 1:**
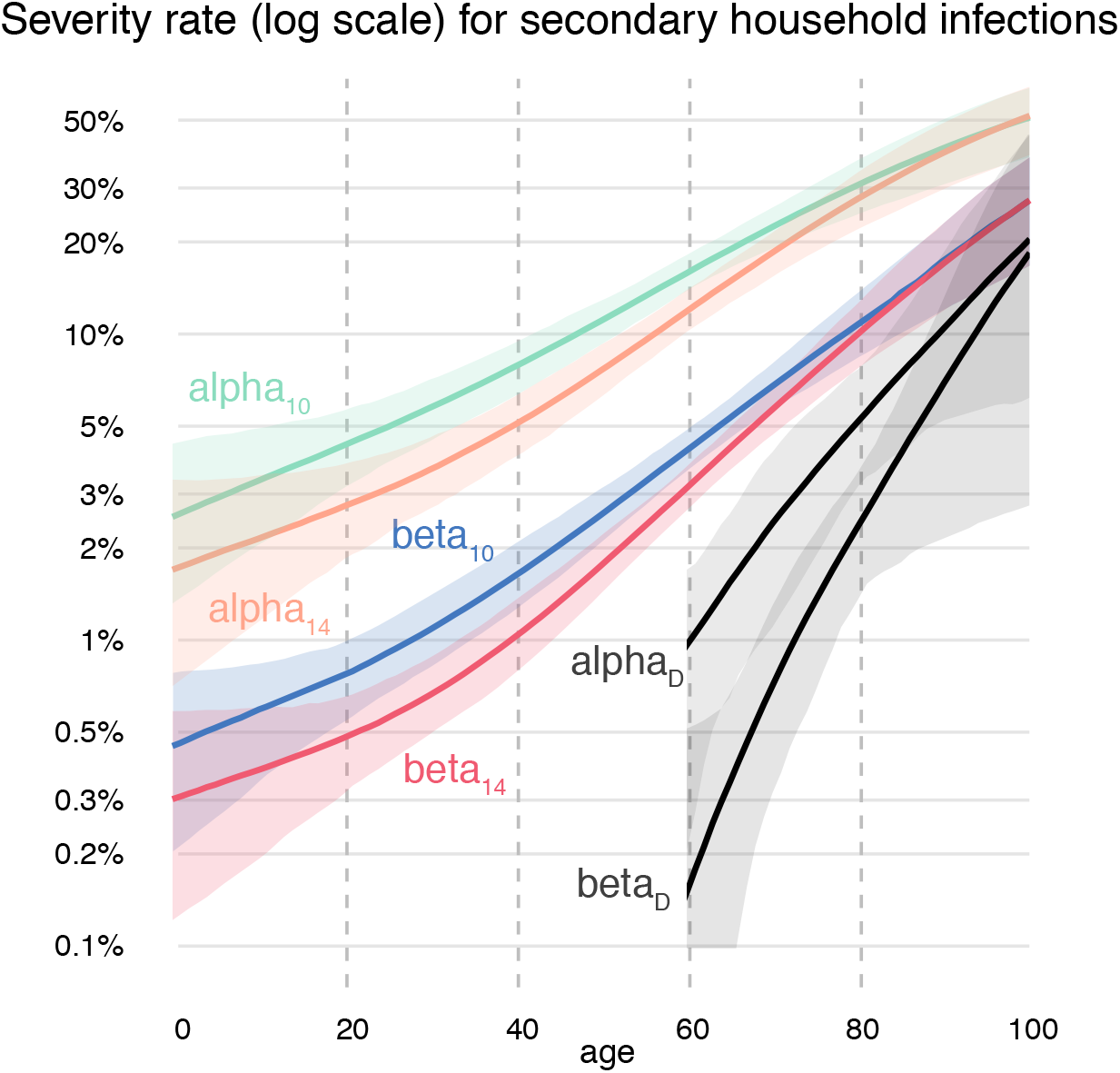
Partial dependence profiles for lower (beta) and upper (alpha) bound for the severity rate and death rate estimated with a linear tail-restricted cubic spline function. Filled regions show 98% bootstrap intervals. The subscript stands for: 10 - severity calculated for 10 days, 14 - severity calculated for 14 days, D - death rate.

### 3.3 Estimation of the bounds on the total number of COVID-19 cases

From the maximum likelihood estimate of the lower bound on the severe case rate *β* and from the number of severe cases *T* ^*sev*^ we obtain the maximum likelihood estimator for the upper bound of the total number of infections 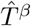 (Equation 5). We found an overall upper bound of 523 796 on COVID-19 infections in Poland (corresponding to 1.38% of the Polish population), using a 10 days threshold for the severe cases (Table 3). This upper bound is around 14 times larger than the cumulative recorded number of cases. The 99 percentile of this upper bound estimator is 660 156 (corresponding to 1.74% of the population). Using the 14 days threshold the upper bound is 585 147 (1.55%) with a 99 percentile of 779 118 (2.06%).

**Table 3:** Estimates of the bounds for the cumulative number of infections in Poland as of July 22nd 2020. Visibility ratio denotes the fraction of detected cases among all cases.

|  | Total | 0-39 | 40-59 | 60-79 | 80+ |
| --- | --- | --- | --- | --- | --- |
| Detected cases in Poland | 41 162 |  |  |  |  |
| Lower and upper bound for the cumulative number of infections<br>in Poland using severeness rate with 10 days threshold |  |  |  |  |  |
| Upper bound estimate $\hat{T}^\beta$ | 523 796 | 251 092 | 164 026 | 92 312 | 16 368 |
| 1-percentile of the upper bound | 423 872 | 200 586 | 134 078 | 76 768 | 12 441 |
| 99-percentile of the upper bound | 660 156 | 320 938 | 204 725 | 112 186 | 22 309 |
| Lower bound estimate $\hat{T}^\alpha$ | 58 746 | 23 127 | 20 346 | 12 523 | 2753 |
| 1-percentile of the lower bound | 48 429 | 18 636 | 16 839 | 10 716 | 2240 |
| 99-percentile of the lower bound | 73 304 | 29 459 | 25 206 | 15 042 | 3599 |
| Visibility ratio | [0.08 – 0.70] |  |  |  |  |
| Lower and upper bound for the cumulative number of infections<br>in Poland using severeness rate with 14 days threshold |  |  |  |  |  |
| Upper bound estimate $\hat{T}^\beta$ | 585 147 | 306 803 | 166 147 | 96 521 | 15 677 |
| 1-percentile of the upper bound | 451 040 | 229 662 | 131 104 | 78 440 | 11 836 |
| 99-percentile of the upper bound | 779 118 | 423 376 | 214 243 | 119 982 | 21 518 |
| Lower bound estimate $\hat{T}^\alpha$ | 64 595 | 28 258 | 20 609 | 13 094 | 2637 |
| 1-percentile of the lower bound | 50 901 | 21 251 | 16 603 | 10 918 | 2131 |
| 99-percentile of the lower bound | 85 301 | 39 163 | 26 592 | 16 074 | 3473 |
| Visibility ratio | [0.07 – 0.64] |  |  |  |  |
| Lower and upper bound for the cumulative number of infections<br>in Poland using death rate only |  |  |  |  |  |
| Upper bound estimate $\hat{T}^\beta$ | 588 853 | | 329 985 | 228 138 | 30 731 |
| 1-percentile of the upper bound | 284 252 |  | 131 461 | 134 881 | 17 911 |
| 99-percentile of the upper bound | 2 118 181 |  | 1 600 389 | 454 261 | 63 532 |
| Lower bound estimate $\hat{T}^\alpha$ | 77 046 | | 40 931 | 30 948 | 5168 |
| 1-percentile of the lower bound | 38 081 |  | 16 527 | 18 413 | 3142 |
| 99-percentile of the lower bound | 270 108 |  | 198 324 | 61 156 | 10 629 |
| Visibility ratio | [0.07 – 0.53] |  |  |  |  |

The lower bound estimate for the total number of COVID-19 cases in Poland 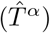 is 64 595 using a 14 days threshold for the severe cases. This lower bound is 1.6 times larger than the recorded number of cases. The 1 percentile of the lower bound was equal to 50 901 and the 99-percentile was equal to 85 301.

### 3.4 Lower bounds on household attack ratio and probability

For this analysis, we split the before considered age group 0 − 39 into 0 − 19 and 20 − 39 and show that attack probability in the 0 − 19 age group was 0.057, around half as high as in the oldest two age groups (0.1273 for the 80+ group). The attack probability estimate for all secondary cases (age-independent) was *λ*^*^ = 0.082. How significantly the data supports the hypothesis that the attack probability is age dependent is discussed in SOM E.

The maximum likelihood estimate for the lower bound on the household attack ratio for the entire set of secondary cases was 11%, while the age dependent estimates ranged from 8.2% for the youngest to 16.9% for the oldest age group, respectively (Table 4).

**Table 4:** Age-dependent attack probability and attack ratio estimation with corresponding known and expected numbers of infected and susceptibles in the age groups.

| Age group | 0-19 | 20-39 | 40-59 | 60-79 | 80+ |
| --- | --- | --- | --- | --- | --- |
| Known secondary infected | 909 | 1259 | 1166 | 694 | 176 |
| Age-dependent attack probability $\lambda$ | | | | | |
| Estimated $\lambda$ | 0.06 | 0.08 | 0.09 | 0.1 | 0.13 |
| Estimated mean number of infected | 919 | 1262 | 1166 | 652 | 177 |
| 99% CI for the number of infected | (839, 999) | (1168, 1360) | (1080, 1252) | (587, 719) | (143, 214) |
| Estimated mean number of susceptibles | 11 238 | 12 301 | 9399 | 5115 | 1047 |
| 99% CI for the number of susceptibles | (10951, 11539) | (12021, 12585) | (9198, 9612) | (4934, 5291) | (961, 1134) |
| Age-dependent attack ratio corresponding to the estimated probability $\lambda$ | | | | | |
| Mean attack ratio | 8.18% | 10.26% | 12.40% | 12.75% | 16.88% |
| 99% CI lower bound | 7.47% | 9.5% | 11.49% | 11.48% | 13.66% |
| 99% CI upper bound | 8.89% | 11.06% | 13.32% | 14.06% | 20.44% |

## 4 Discussion

We propose a new method for the estimation of severeness and attack rates, as well as the unknown total number of SARS-CoV-2 infections, including diagnosed and undiagnosed cases. The estimation is based on routine surveillance data, i.e. the residence address of the case and the indication of severity of the disease course. In contrast to approaches based on seroprevalence testing, it does not require expensive population studies. Compared to model-based approaches, it is data-based and introduces only minimal assumptions.

The assumption that severe cases or deaths are likely to be diagnosed and registered was made previously by other authors, e.g., Flaxman et al.^17^ These approaches, however, were limited by the lack of precise information on the expected infection fatality rate or, more generally, the expected fraction of the severe cases. The assumption that secondary infections in households constitute an unbiased sample of the infected, with similar severeness rate to the entire population, was considered also by Hernandez-Suarez et al.^18^ This work, however, relied on asymptotic approximations, did not account for problems encountered in real data, such as unknown household sizes, and finally, did not apply their method to any case dataset.

We estimate that from 7% up to 70% of all cases in Poland were actually detected. As at the time of the study the strategy was to test all quarantined individuals, we consider higher detection rates and thus also the lower bound estimates of the total prevalence to be the more likely. The low estimated prevalence bounds are in agreement with the seroprevalence below 1% measured in a population sample in Czechia,^19^ which has a similar detection rate as in Poland. Up to date, there are no published seroprevalence results in Poland that we could compare the estimates to. However, a study in Cracow reports 2% seroprevalence in this city [personal communication K. Pyrć]. Importantly, even the unlikely upper prevalence bound estimate 2.06% of the whole population (for severity defined as 14 days) is not enough to provide any effect of herd immunity.^20,21^

To our knowledge, we are the first to derive the household attack probability, also in different age groups, and show how the attack ratio depends on this quantity. Since the household attack ratios depend on household distribution, it is recommendable to report attack probability instead, which is rather an intrinsic parameter of the disease.

The approach has several limitations. The results depend on the definition of the severe case and how accurately the severe cases are recorded in the data. On top of that, precise recording of the household sizes and their age distribution would lower the variance of our estimations.

In conclusion, the method is easily applicable using surveillance data and provides useful information on the severeness and attack rates, as well as the total number of infections and the undiagnosed fraction. In the future, it could be used to continuously monitor the effectiveness of the testing strategy and the proportion of individuals who have already passed the infection. In the example of Poland we show that only a minor part of individuals were already infected and recovered.

## 5 Ethics approval

The protocol was approved by the Bioethics Committee of the Medical University in Wroclaw (KB–610/2020) and complies with the Declaration of Helsinki of the World Medical Association.

## Data Availability

All data generated or analysed during this study are included in this published article.

## 6 Acknowledgments

The MOCOS group thanks the Polish National Institute of Public Health and the City of Wroclaw for financial and logistic support, as well as the Central Statistical Office of Poland for providing access to the 2011 census data.

## Supplementary online material

### A Smooth version of the severity rate estimator

In equation 2 the rate *α* is defined as the expected rate of severe infection among household secondary infections. This rate may be dependent on some observed characteristics of infected persons, like age, sex or comorbidities. Let *α*(*x*) be an expected severeness rate for an individual with observed characteristics *x*. For simplicity, we have considered only the most important characteristic i.e. *age*, but the approach works also for more general settings.

Let 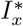 be the number of observed secondary infected individuals with observed characteristic *x*. Assuming that these infections are independent the number of observed severe cases 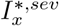 is a binomial random variable

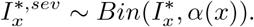

If groups 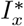 are large enough then we can estimate *α*(*x*) in each group independently with the procedure described in equation 9. Such estimates for four age groups are presented for example in Table 2.

If we want to have a continuous form of the function *α*(*x*) then we can treat this problem in the same way as in the classification problem. We assumed that *α*(*x*) can be approximated by some family of functions. A simple logistic regression with a linear link may be too rigid. We compared the gradient boosting approach and logistic regression with restricted cubic splines [22]. Both leads to similar results thus only the one with splines, as more smooth, is presented below.

For age, we used four knots places in percentiles 5, 35, 65, 95. This corresponds to age breaks at 14, 40, 56, 83. Between knots, the function *α*(*x*) is approximated as cubic polynomial while outside knots it is approximated as a linear function. Additional restrictions are put to get smooth approximation in knots.

The exact formula of three cubic polynomials is hard to read so to visualise this relation we used the Partial Dependence profiles implemented in the DALEX [23] library for R [24]. The relation is presented in Figure 1. Note that due to the behaviour of *α*(*x*) and *β*(*x*), the log-linear axes are used in the plot.

The procedure for the *β*(*x*) is similar, with the only difference that instead of the number of observed infected cases 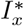 we use 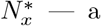; — a census-based estimation of the size of the maximum susceptible population size with characteristics *x*. See appendix D for details.

The 98% pointwise confidence intervals presented in 1 is obtained with the bootstrap procedure based on 1000 bootstrap samples. In each bootstrap sample, the households were sampled with replacement and used for estimation of *α*(*x*) and *β*(*x*).

These results can be reproduced with scripts available at the https://github.com/MOCOS-COVID19/dark-figure.

### B Derivation of the one-sided confidence bounds on severeness rates and the total number of infections

A conservative approximation to the confidence interval bounds is obtained by the Clopper-Pearson interval.^12^ To obtain an upper bound estimator of infected we need the one-sided q% lower confidence interval bound of *β*. This is obtained by finding the value *<u>β_q_</u>* = *θ* ∈ [0, 1] with *P* (*x* ≤ *I*^*,sev^) = *q* where *x* ∼ *Bin*(*N*^*^, *θ*). Therefore the one-sided q% upper confidence interval bound for the upper bound of infected is given by

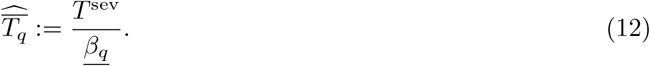

In the case when the household size distribution is unknown, we extend the idea of the Clopper-Pearson interval by searching for the value of *<u>β</u>*<u>_*q*_</u> = *θ* ∈ [0, 1] with 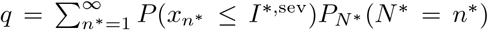 where 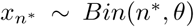. Since 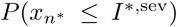 is monotonic in *θ* for each *n*^*^ also 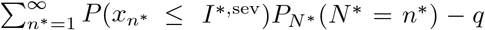 is monotonic as a convex combination. We then use the bisection method to find the single root of this function.

Analogously, the one-sided q% lower confidence interval bound for the lower bound estimator of infected can be derived by obtaining the 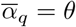 with *P* (*x* ≥ *I*^*,sev^) = *q*, where *x* ∼ *Bin*(*I*^*^, *θ*)

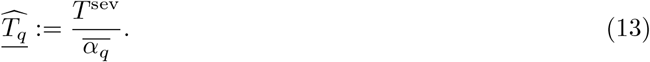

In the case without observed severe cases in the secondary infections, the rule of three by Eypasch et al. can be used to approximate a 95% confidence interval bound instead.^25^

### C Adjusting for delayed *T*^sev^

Let *p*(*k*) be the probability of developing a severe progression after *k* days after infection, conditionally on developing the severe progression at some point in time. Denote as before by 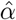 the severe case rate and let *ϕ*(*t*) be the cumulative number of severe cases discovered at day *t*. Further, we denote by *δ*(*t*) be the number of new severe cases manifesting themselves at day *t*, and by Δ(*t*) the number of all new infections (not only the discovered) at day *t*. For the following considerations we assume that the daily reported number of severe progressions is reported without delay. However, if there is a delay in reporting, then the estimated number of infected have to be shifted backwards by this delay.

In particular, for the *k*-th day the amount of new severe cases among the previously infected is

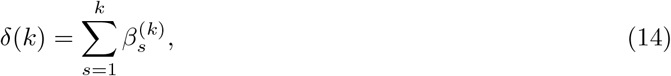

where the 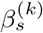 are realizations from the Binomial distribution given by 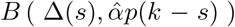. If the immunity is not complete and a second infection is possible, these probabilities may change for the second infection. Then the probability of developing a severe progression in the second infection may be lower, and the estimated figures will tend to underestimate the total amount of infected.

At day *s*, Δ(*s*) persons get infected. The probability to exhibit a severe progression at day *k* conditionally on developing a severe progression at some point is *p*(*k* − *s*). In expectation the number of severe cases at day *t*, starting at day *T*_0_, is then given by

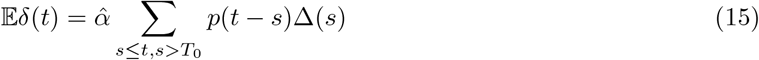

Recall that *δ*(*k*) is a sum of independent Bernoulli random variables. Define *μ* := 𝔼*δ*(*k*). By applying the Chernoff bound (see e.g. Theorems 4.4 and 4.5 in Mitzenmacher and Upfal^26^), we get for *λ* > 0

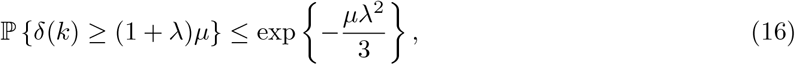

and

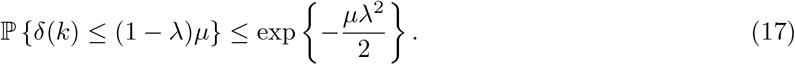

For a single realization *r* of *δ*(*k*) we get

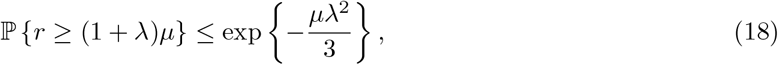

and

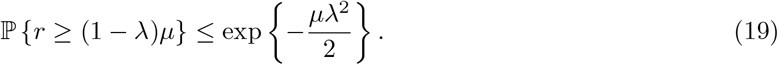

For obtaining a lower *Q*-significant estimate on *μ* from (18), we take *λ* = *r/μ* − 1, that is *r* = (1 + *λ*)*μ*, and find the solution to the constrained optimization problem

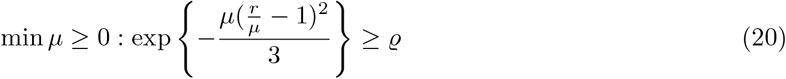

The solution to (20) is the smaller root of the equation with unknown *μ*

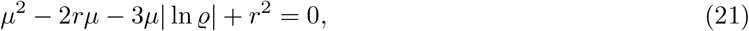

so 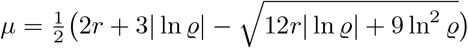.

Similarly, by taking *λ* = 1 − *r/μ* in (19) we can find an upper *ϱ*-significant estimate on *μ* by solving the constrained optimization problem

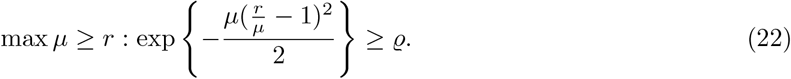

The solution to (22) is the greater root of

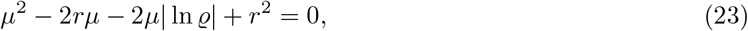

hence so 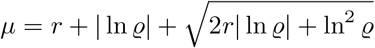.

Take now 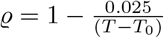, and let *r*_*k*_ be the realizations of *δ*(*k*) that we observe. Set

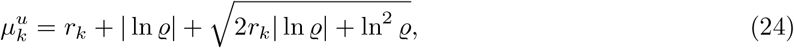

and

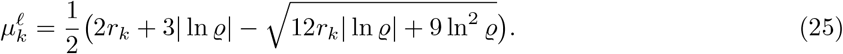

Then by (14) and (15)

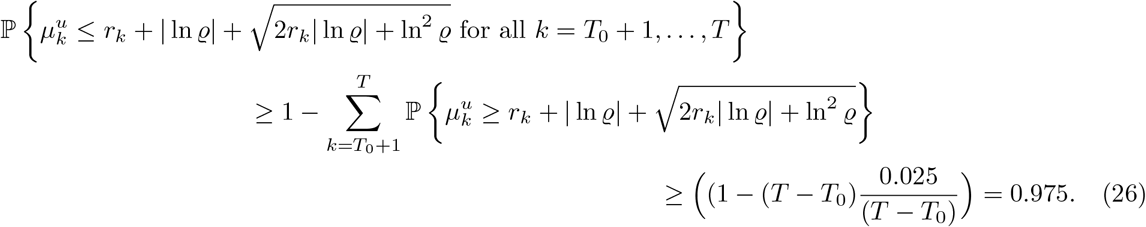

Hence 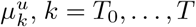, defined by (24) give us upper 0.025-significant estimates for Δ(*k*), *k* = *T*_0_, …, *T*, in the sense that, assuming 𝔼*δ*(*k*) = *μ*_*k*_,

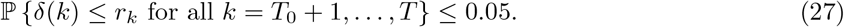

Similarly, 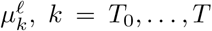, defined by (25) provide lower 0.025-significant estimates for Δ(*k*), *k* = *T*_0_, …, *T*.

We can rewrite (15) as

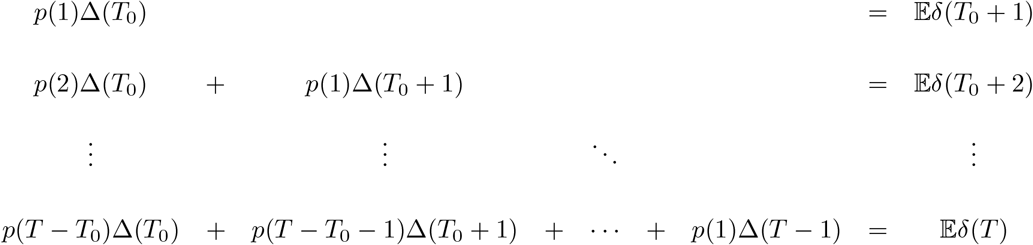

We see that the coefficient matrix is a triangular matrix and thus the system can be solved by forward substitution. In particular,

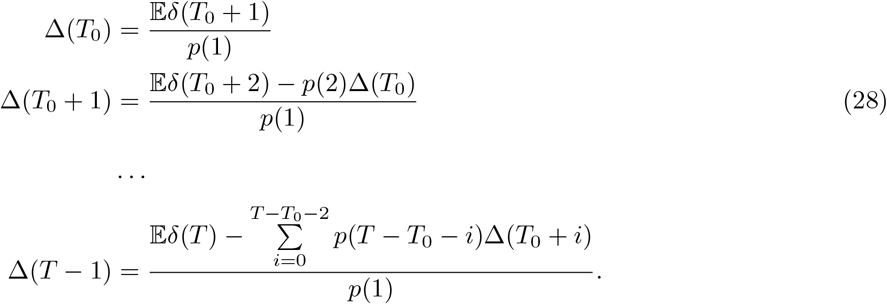

Denote by 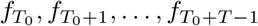 the solutions to (28) as linear functions of 𝔼*δ*(*T*_0_ + 1), 𝔼*δ*(*T*_0_ + 2), …, 𝔼*δ*(*T*).

The lower and upper boundaries of a 0.05-significant confidence interval for Δ(*t*), *t* ∈ {*T*_0_, *T*_0_ + 1, …} are given by

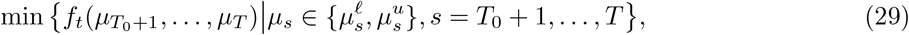

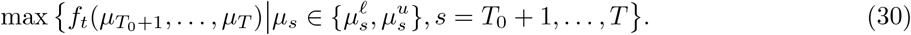

Similarly, the cumulative number of infected 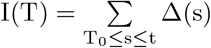 can also be expressed as a linear function *F* of 𝔼*δ*(*T*_0_ + 1), 𝔼*δ*(*T*_0_ + 2), …, 𝔼*δ*(*T*), hence the boundaries of a 0.05-significant confidence interval are

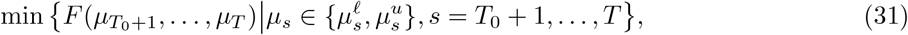

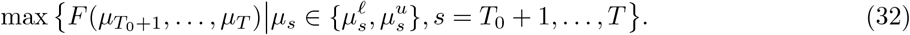

#### Remark

From the numerical point of view the method outlined here works well if 1 is the mode of the distribution *p*, or at least if *p*(1) is close to the max{*p*(*k*)}. Otherwise the elements of the inverse matrix may take very large values, and dependence of Δ on *δ* is very sensitive, making the approach not numerically stable. Additionally, if the aim is to obtain a confidence interval only for the total number of infected I(T), a narrower confidence interval can be designed.

### D Estimation of the susceptible population size *N*^*^ under unknown household sizes

Unfortunately, the information on the household size of the index patients has not been recorded in Poland and thus has to be estimated. For the estimation we used the data from 2011 Census.^27^ A representative study was done on a random sample of approx. 20% (approx. 2 744 000) households in Poland, out of the total number of 13.5 million registered households. The data was successfully collected directly from inhabitants of 2 272 711 households.^28^

Based on the data described above, we estimate the average household size to be 3.35. A (1 − *ϱ*)100% confidence interval can be obtained using Hoeffding’s concentration inequality in the form 3.35 *± q*_*ϱ*_, where *q*_*ϱ*_ solves 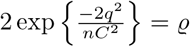with *C* = 56 being the maximum household size. In particular, a 99% confidence interval is 3.35 *±* 0.0605, and a 95% confidence interval is 3.35 *±* 0.0505.

In Figure 2 we give the distribution of mean household size given the age of a randomly chosen individual along with the standard deviation in the population of Poland. Note, that we use the household size in terms of the number of people living together. This is in contrast to the majority of publication defining the household as economic unit. For the economical unit the mean household size was 2.98 in the census 2011.

**Figure 2:**
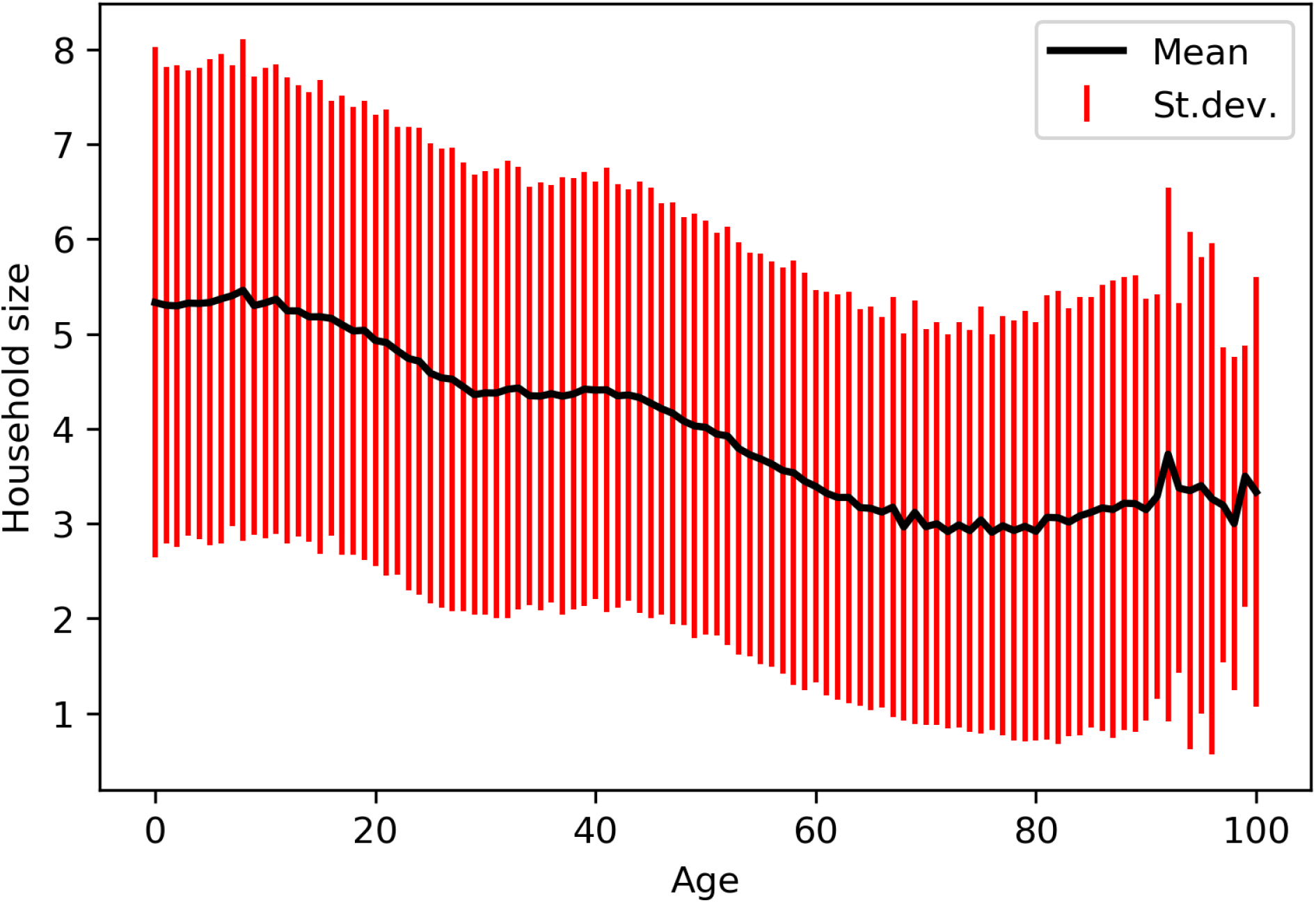
Mean and standard deviation of household size given the age of a randomly chosen individual from the population of Poland.

#### D.1 Estimation of household size

For each index case, we sampled a household *h* from “National Census of Population and Housing 2011” inhabited by a person of the same age *a* and sex as the index case, and calculated the number of household members within each considered age group *g* in *G* = {0 − 39, 40 − 59, 60 − 79, 80 + years old}. For each household *h* we hence obtained 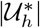 as the sum of the number of household members in all age groups 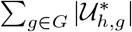. This bootstrapping procedure was repeated 10000 times. In each iteration *w* of the procedure, after all index cases had been processed, the numbers of household members in each age group were totalled, 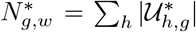. We estimate the size of the susceptible population in each age group 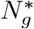 as the 99th percentile of all obtained 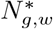, and the total size of the susceptible population as 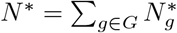.

#### D.2 Estimation of the household size with partial information on the household size

The above procedure yields mean household sizes when no household size information is known, i.e. under the assumption that for each *h* the total household size 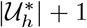 is at least 1. However, in cases when other household members, but possibly not all of them, were infected and these links were reported in the SRWE data, we are able to determine the minimal size of these particular households. Based on the SRWE data, we calculated a minimal household size *k* + 1 for each index case as the number of all infected people living in the same household. Thus, for an index case of known age *a*, sex, and minimal household size *k* + 1 we sampled only from households satisfying all three conditions. For people younger than 18 years old, who legally cannot live alone, we set max(*k* + 1, 2).

#### D.3 Estimation of the household size including spatial data

In the SRWE database, the exact address of the residence of each case is reported. Since household size distributions may vary across voyvodships and, moreover, the distribution of household sizes at the voyvodeship level is available in the 2011 Census data, we included the voyvodship information in the bootstrap procedure. Thus, for each index case, in addition to the age, sex, and the minimal household size conditions, we conditioned the household sample on a voyvodship. The only exception from this condition was made for Podlaskie voyvodship, for which very few households are provided in the 2011 Census data. For this particular voyvodship we used the household size distribution of Poland.

Figure 3 illustrates the differences in the distributions of the number of susceptibles in each age group obtained by the three bootstrapping procedures mentioned above. The procedure that served as a basis for the results presented in Table 2 and Table 3, was the third procedure that takes into account both the minimal household size and spatial data, due to its richest usage of available data.

**Figure 3:**
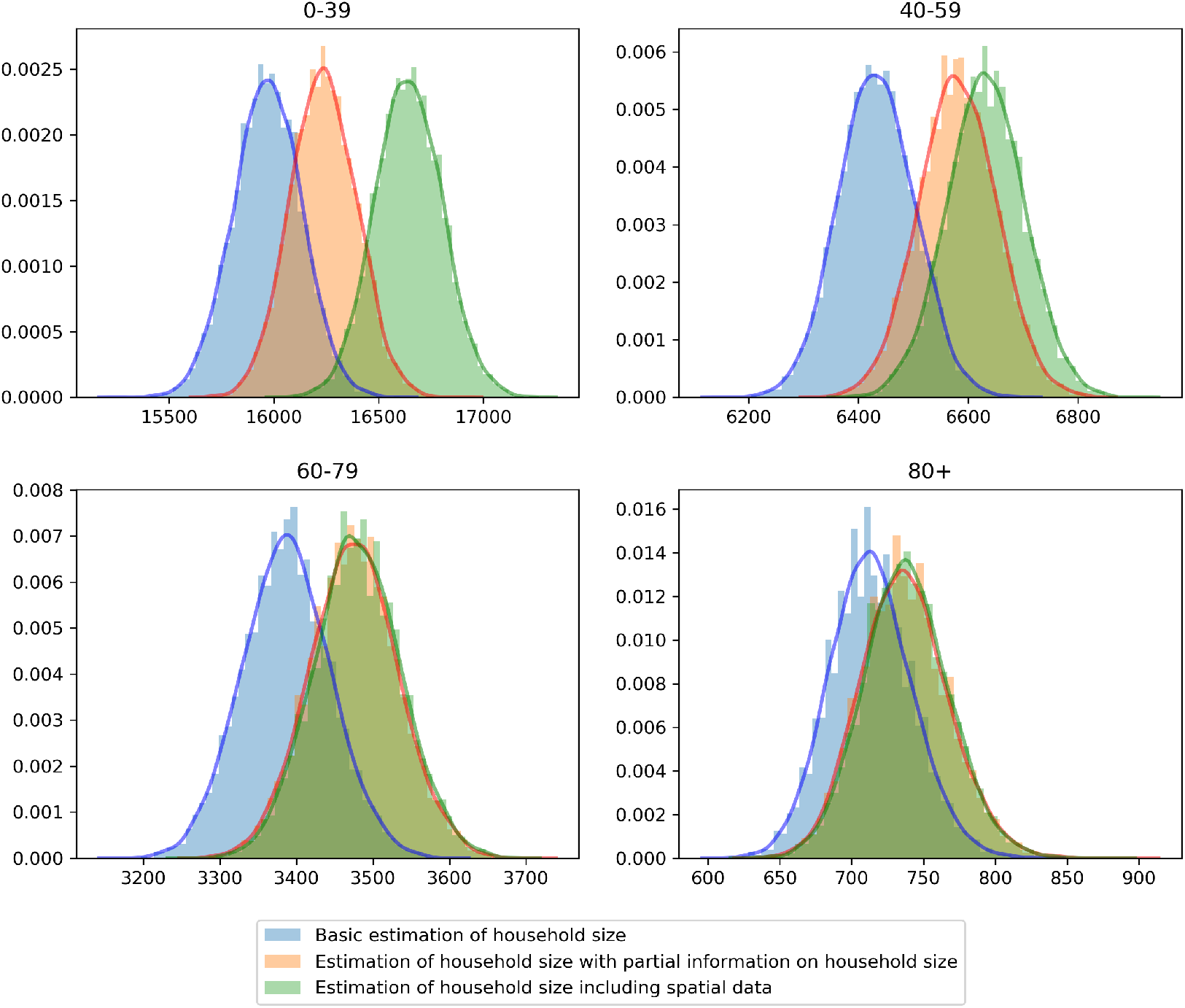
Distribution of the number of susceptibles in each age group based on the results of the bootstrapping procedure.

In Figure 4 we compare empirical cumulative distribution functions (ECDF) of age within reported index cases and secondary case to the ECDF of age within the general population of Poland. The index cases population is clearly older than the general population, whereas the distribution of age of secondary cases resembles the distribution of age in the general population. Further, the distribution of age within susceptible population, obtained from the bootstrapping procedure, indicates that this population is younger than the general population.

**Figure 4:**
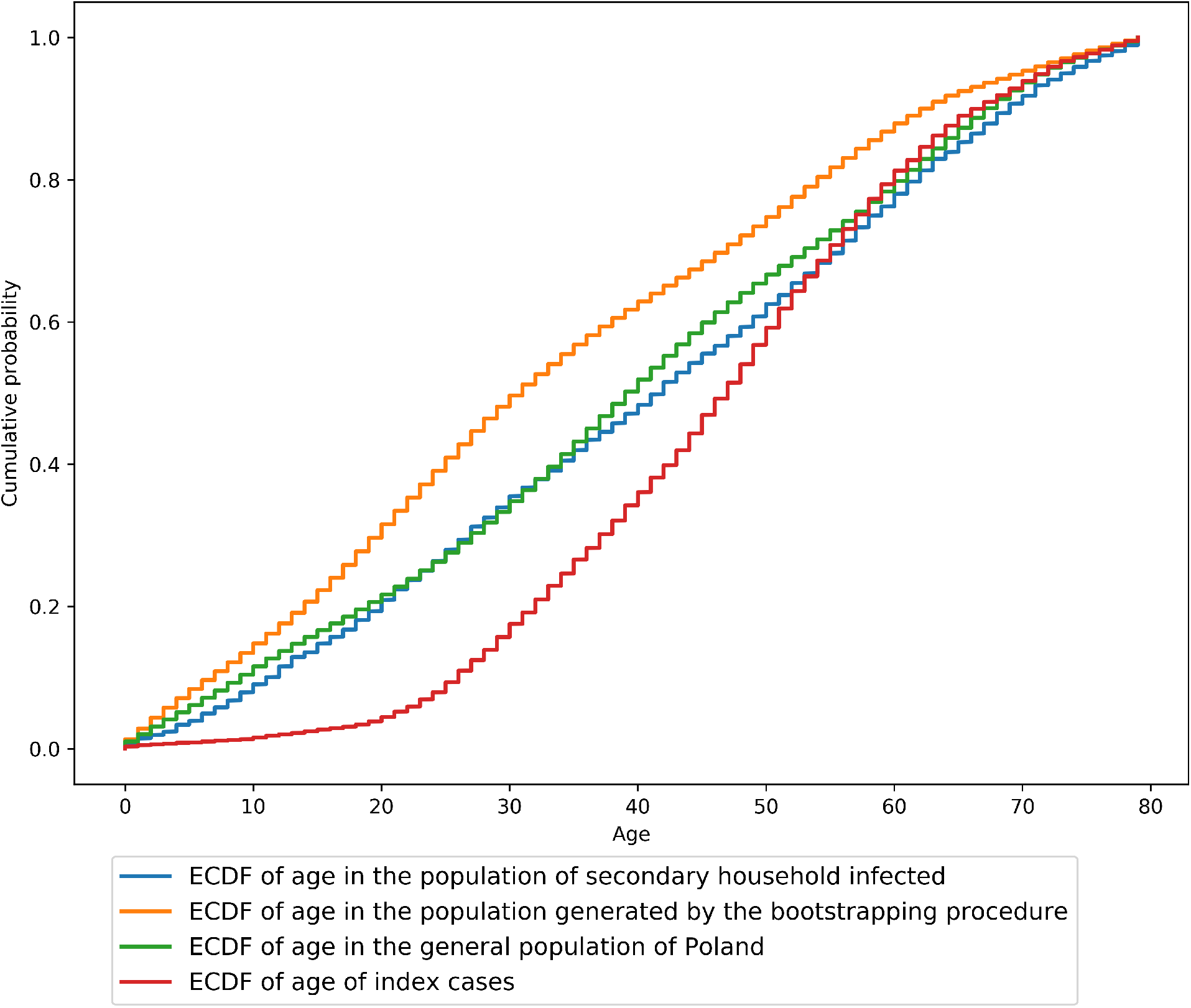
Empirical cumulative distribution functions of age in the population of secondary household infected (based on the SRWE data), in the population generated by the bootstrapping procedure (based on 2011 Census data), in the general population of Poland (based on official 06.2019 statistics), and of index cases (from SRWE data). All four ECDFs illustrate the cumulative probability of age among people younger than 80 years old.

In Figure 5, the frequencies of household sizes of an infected population are given. In case the source of infection is known and the circumstances are classified as “Household contact” we make the assumption that all household members were infected and use this assumption to calculate their household size. In case the source of infection is from the outside of the household or is unknown, then we take the average household size given the age of a person.

**Figure 5:**
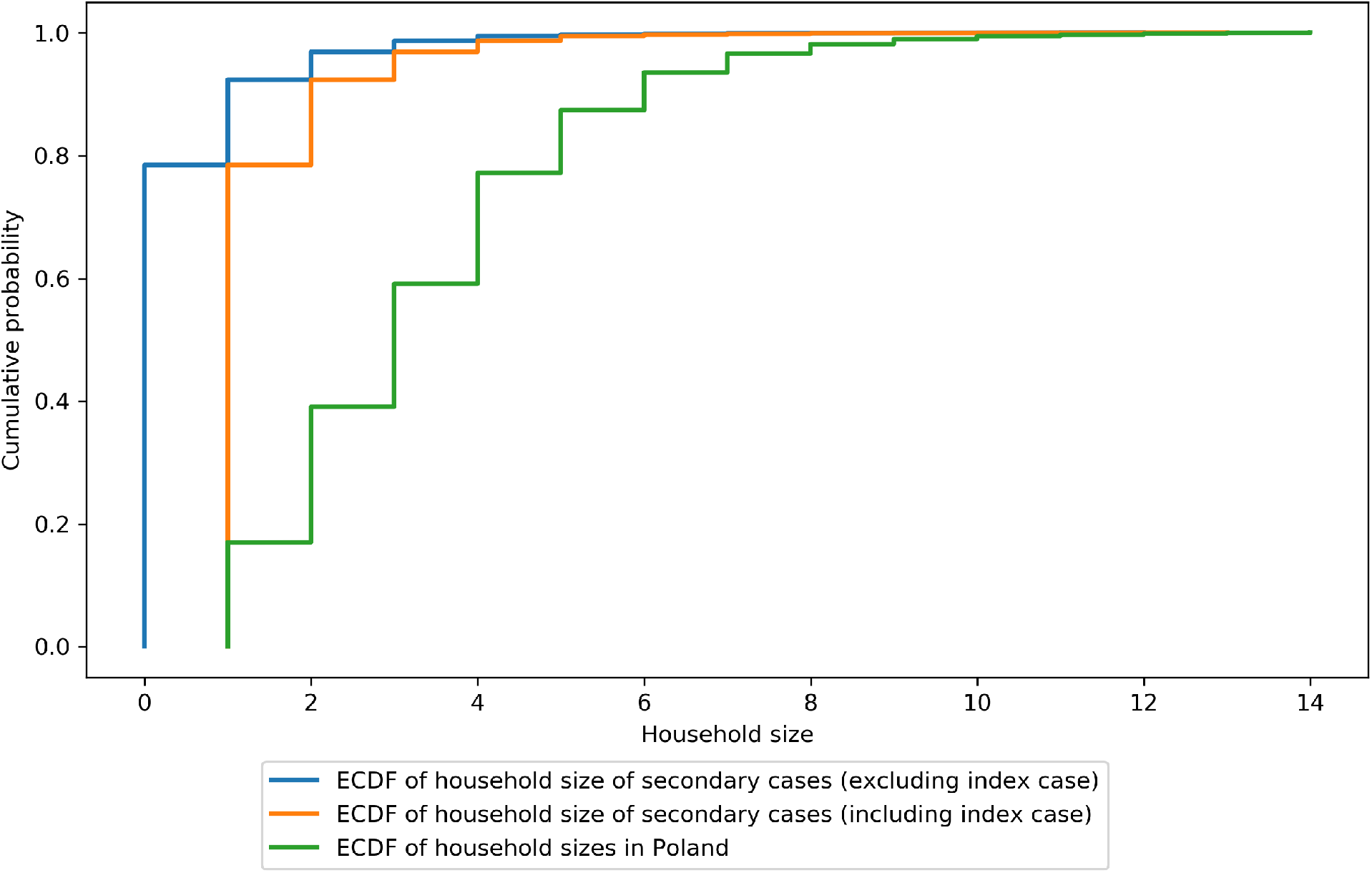
ECDF of household sizes (with less than 15 inhabitants) in the general population of Poland and within the population of secondary cases. In case the source of infection is known and the circumstances are classified as “Household contact” we make the assumption that all household members were infected and use this assumption to calculate their household size. In case the source of infection is from the outside of the household or is unknown, then we take the average household size given the age of a person.

### E Estimating lower bounds on the in-household attack rate *λ*

We describe here in detail how to get a lower bound on the attack rate within a given age cohort. Assume we have *n* index patients enumerated from 1 to *n*. For the attack rate we will use as index patients the first infected patient in a household, that is the patient whose source of infection was outside the household or the patient where the source is not known. We assume a constant attack rate *λ* ∈ [0, 1] which is defined as the a priori probability of an index patient to infect a given member of the household. Let *H*_*i*_ be the sampled household of index patient *i* and let the random variable *h*_*i*_ be the number of susceptibles in household *H*_*i*_. We first discuss the situation when *λ* does not depend on the age of the index patient nor on the age of the susceptibles in *H*_*i*_. To link the attack rate with the observed number of cases in the susceptible secondary household population we need first to estimate the expected number of infected in a household of given size. Let *μ*_*k*_ (*λ*) be expected number of infected in a household with susceptible size *k* (not counting the index patient). Trivially we have *μ*_*k*_ (*λ*) ≥ *λk*. Let *Y*_*i*_ be the random variable of the actual (unknown) number of secondary infections in household *H*_*i*_. We consider only households up to size 15. Then the actual number *I* of infected in the susceptible population is given by

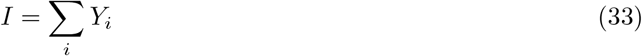

where the random variables *Y*_*i*_ are independent but not identical distributed. Due to the concentration properties of sums of bounded independent random variables, *I* is concentrated around the expectation 𝔼*I*. Since the household-size distribution depends on age we have to group the index patients into age classes [*a*] corresponding to the age cohort *a*. Clearly

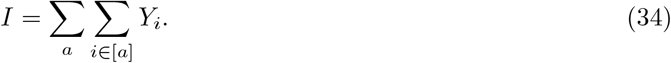

Let further *p*_*k*_ (*a*) be the probability that an index patient from age class [*a*] lives in a household of size *k* + 1. For *i* ∈ [*a*] we have

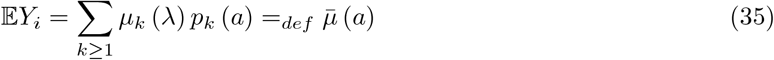

where 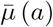 is the expected number of secondary household infected for index patients in a age class [*a*]. We have finally

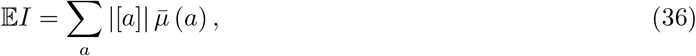

where |[*a*]| is the number of patients in the cohort [*a*]. Let further *N*^*^ be the total number of the susceptible population, that is

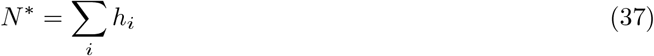

and

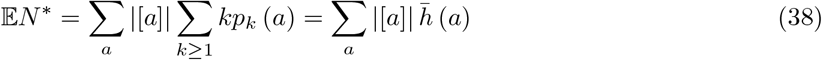

where 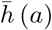 is the expected secondary household-size of an index patient in class |[*a*]|. By the law of large numbers we have for large numbers of index patients

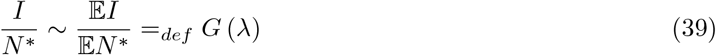

(we can make the errors explicitly by using concentration inequalities). Clearly *G* (*λ*) - the fraction of true case in the secondary household population - is a continuous and strictly monotone increasing function in *λ* and has an inverse. In Figure 6 we give the obtained *G* (*λ*) for *λ* ∈ [0, 0.25].

**Figure 6:**
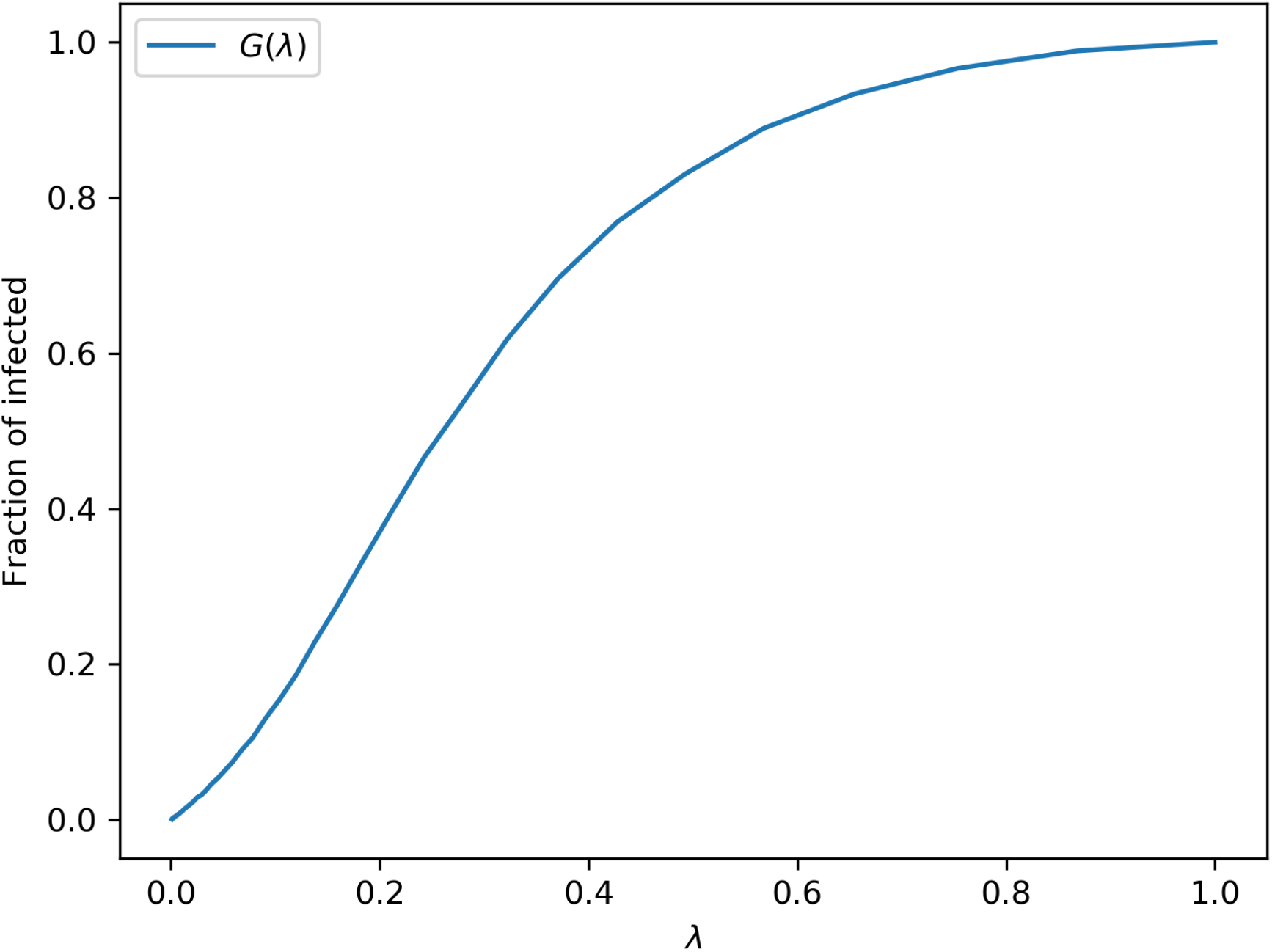
Fraction of infected within secondary household members

Given the observed number of secondary household infections *Î* we get under the assumption that *Î* is the true number of cases as an estimator for the attack rate

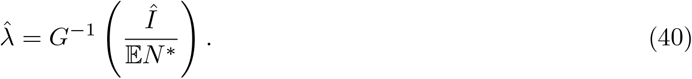

Furthermore 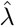 defines a lower bound on the true attack rate since we assumed that *Î* is the the true number of cases.

Note that there is a close connection to the upper and lower bounds on the severeness rate estimated in the main text. The lower bound severeness probabilities (*β*) were obtained by assuming that all of the susceptible population *N*^*^ is infected, which corresponds to the case *λ* = 1. The upper bound on the severeness rate was obtained by assuming that the observed number of cases *Î* is the true number of cases, hence this corresponds to the attack rate 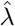.

On the other side, if the true value of 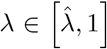 of the attack rate would be known and is independent of age we could estimate the severeness rate *τ* (*a*) in age group *a* as follows. The number of cases *I* (*a*) in age group *a* is given by

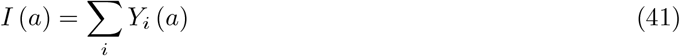

where *Y*_*i*_ (*a*) is the number of infected in households *H*_*i*_ in age group *a*. Let *ν*_*k*_ (*a, b*) be the expected fraction of secondary household members of age class *a* in an household size *k* of an index patient *i* from age class *b*. Then

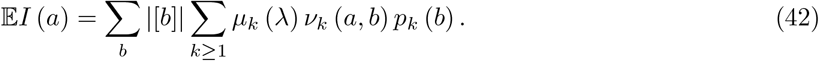

Note that the age classes for *a* and *b* in the above formulas need not to be the same (usually we take the age class *b* for the index patients to consist of a single year, whereas the *a* cohorts are taken to be much larger). Again by the law of large numbers 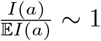 and the maximum likelihood severeness rate based on an observed number *Î*_*sev*_ (*a*) of severe cases in age class *a* reads as

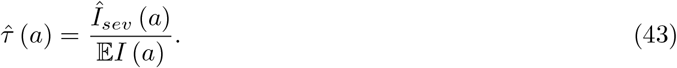

The numbers *ν*_*k*_ (*a, b*) can be computed with arbitrary precision by bootstrapping from the census household population.

The adaptation of the above consideration to the case of age dependent attack rates is straightforward.

#### E.1 Age-dependent attack rates

We first discuss the situation when the attack rate depends on the age of the susceptible but not on the age of the index patient (that is the source of the infection in the household). Let *K* be a partition the age classes of the the susceptibles into *k* age cohorts. For the index patients we assume usually a finer partition *A* into age classes (usually one class per year). Let *λ* = (*λ*_1_,*λ*_2_ …, *λ*_*k*_) ∈[0,1]^*k*^ be the vector of attack rates for the different age cohorts from *K*. Let 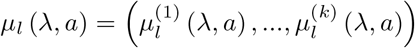 be the vector of expectations of the number of infected in the different age classes in a household of size *l* + 1 conditioned that the index patient is of class *a*. Note that the expectation has to be taken over all possible age compositions of the households. Let finally for each index patient *i*, 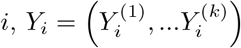 be the vector of the numbers of infected household-members in *H*_*i*_ in the corresponding age classes from *K* and let *I* = (*I*_1_, …, *I*_*k*_) be the vector of numbers of infected in the different age classes in the whole susceptible population *N*^*^. We get in complete analogy with the age independent case

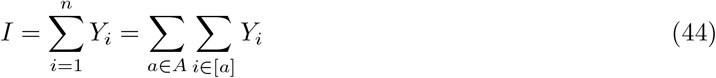

and for the expectation

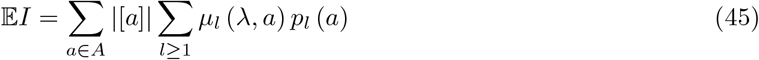

where *p*_*l*_ (*a*) as before is the probability that an index patient of age *a* lives in a household of size *l* + 1. Contrary to the age independent case, *μ*_*l*_ (*λ, a*) might depend on the age *a* of the index patient since the age composition of households matters here. Again by the multivariate law of large numbers we have

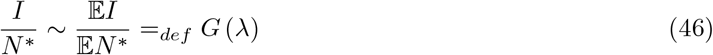

where *G* : [0, 1]^*k*^ → [0, 1]^*k*^ is a strictly monotone increasing mapping and hence as a well defined inverse. Given the vector *Î* = *Î* _1_, …, *Î* _*k*_ of observed infected in the different age groups and in the susceptible population. We get the estimation

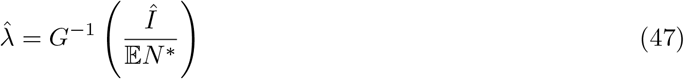

for the attack rates in the age groups from *K*. Instead of the normalization by 𝔼*N*^*^ one could also scale the different components of *I* respectively *Î* by the expected size 𝔼*N*^*^ (*b*) of the susceptible population in an age cohort *b*. This might look from an epidemiological point of view more natural but would only change the definition and form of *G* and give in the end the same (at least asymptotically) estimator 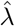. Note that there is no easy analytic way to compute the values of *μ*_*l*_ (*λ, a*). Here one has to rely on numerical approximations for instance by Monte Carlo simulations. The case when the attack rate depends also on the age of an index patient is more complicated and will be discussed elsewhere.

#### E.2 Estimating the age-dependent attack rate from data

We consider attack rate for five age groups: 0-19, 20-39, 40-59, 60-79, and 80+. Thus, *λ* is a vector in five-dimensional space, where each dimension corresponds to a respective age group. We start with a search interval of [*λ*_*l*,0_, *λ*_*u*,0_]^5^ = [0.05, 0.15]^5^. The search is performed by repeatedly bisecting the hypercube defined by these values and then selecting the sub-hypercube in which the bootstrapped mean number of infected equals the observed number of infected within each age group. The algorithm begins with drawing a household for each index case from the 2011 Census data so that age, sex, minimal household size and voyvodship matches those of the index case; the household selection is repeated 10000 times. In the next step, we perform 6 iterations of hypercube bisection and in each iteration we take the center of a current sub-hypercube as a *λ* vector.

For each household configuration, i.e. the vector of the number of susceptibles within each age group in this household, and for the current *λ* vector we calculate the probabilities of infection for each age group and “infect” household members accordingly. For each age group we check whether the mean number of bootstrapped infected is higher or lower than the observed number of secondary infected within this age group and, depending on the result, the next iteration is performed on an interval [*λ*_*l,i*_, *λ*_*i*_] or [*λ*_*i*_, *λ*_*l,i*_], respectively.

#### E.3 Discussion of uniform versus age-dependent attack rate

We further attempted to test the hypothesis that the attack probability is age independent. To this end, we compared the numbers of known infected individuals to the numbers expected given the lower bound uniform attack probability *λ*^*^. By the definition of the attack ratio, the expected total number of infected individuals should depend on the attack ratio and on the expected household size as

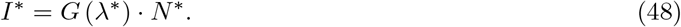

In the case when the *λ*^*^ was indeed age independent, the recorded numbers of infected individuals in the corresponding age groups should at least be inside the 99 percent confidence bounds of with generated number of secondary infections (based on the estimated uniform attack probability).

The expected numbers of infected individuals in different age groups with their 99% confidence bounds, obtained for the estimated uniform, age-independent *λ*^*^ = 0.082, computed according to the estimation procedure described in Section E.1 are presented in Table 5. In the youngest age cohort (0 − 39 years) the actually recorded number of infections is smaller than the lower bound of the 99% confidence interval for the number of infected, while for the age group above 40 years old the recorded numbers are above the confidence bounds. Hence, we would reject the hypothesis that the attack rate does not depend on age if we could be sure that the observed number of cases *I*^*^ is really the true number of cases. To illustrate the age dependence of simulated versus recorded number of infections - assuming a uniform attack probability - we show additionally in Fig. 7 the cumulative empirical distribution function for secondary household infections as a function of age. Finally we still give a uniform lower bound on the attack probability. To get such bound we estimated the largest uniform attack probability *λ*^*^ such that all observed numbers of infections in the corresponding age groups are larger or equal to the 99 percent upper bound values of expected infections given the value of *λ*^*^. We estimate this *λ*^*^ to be 0.0576 and the corresponding values of secondary infections are given in table 6.

**Table 5:**
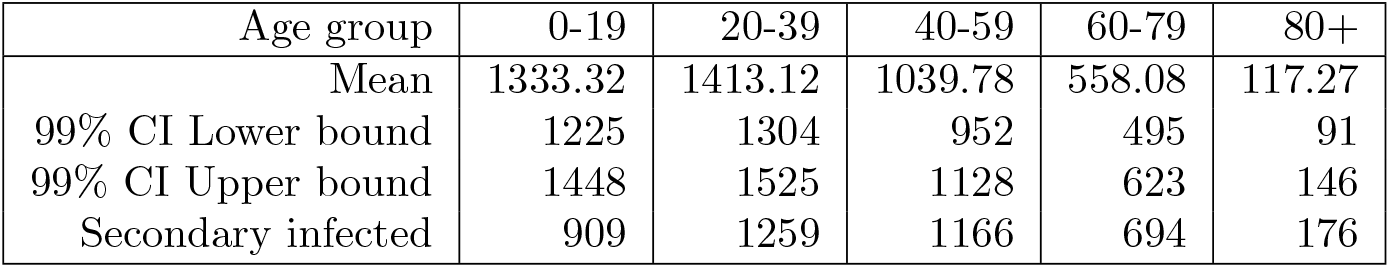
Expected number of infected assuming a uniform attack probability equal to 0.082

**Table 6:**
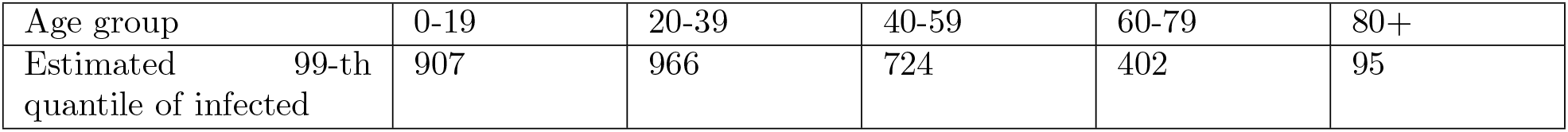
Expected 99% quantile of the number of infected assuming a uniform attack probability equal to 0.0576

**Figure 7:**
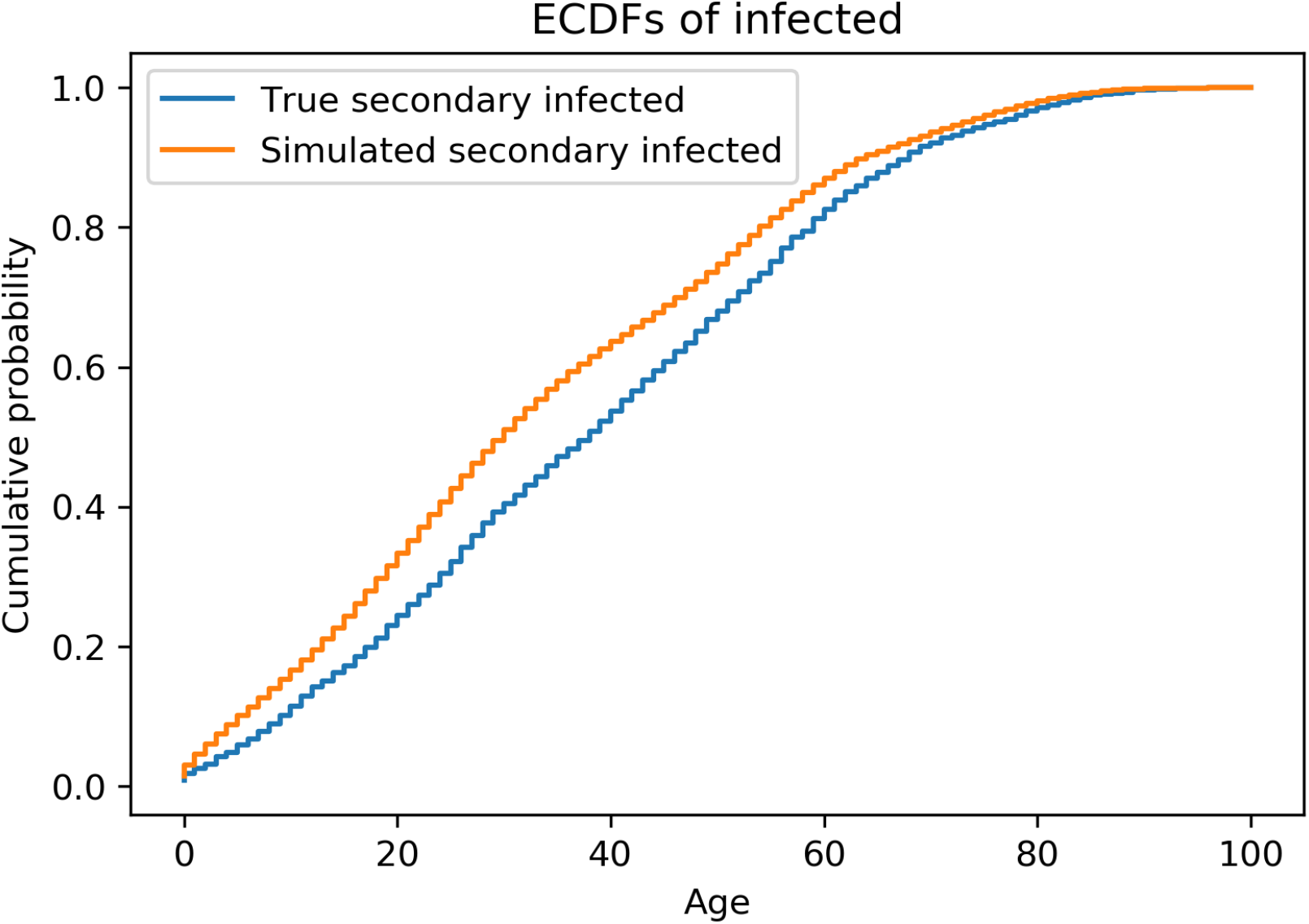
Empirical cumulative distribution functions of true secondary infected vs simulated secondary infected

### F Pre-processing of the surveillance data

First, we extracted the case clusters of size at least two with documented household transmission (the *infected households*). Only cases for which clear epidemiological links were registered as household transmission together with their source cases were included. Cases in social care units and households of minimum 15 inhabitants were filtered out, as an initial analysis revealed that those were not representative for the overall population, due to over-represented comorbidities and severe cases. This filtering left 16 123 cases (summarized in Table 1). In each infected household, the index case was identified as the one with the earliest date of diagnosis, since this case was the most likely to trigger the contact tracing. Other cases in each of the infected households were regarded as secondary cases and included in the estimation of the severe case rate.

